# Bench-stepping training improves stair-walking dynamics in older women: evidence from an exploratory nonlinear kinematic analysis

**DOI:** 10.64898/2026.07.02.26357116

**Authors:** Remco J Baggen, Kimberley S van Schooten, Evelien Van Roie, Sabine M Verschueren, Christophe Delecluse, Kim Delbaere, Stephen R Lord, Jaap H van Dieën

## Abstract

**Introduction:** Stair walking challenges balance and coordination in older people. Bench-stepping training improves stair climbing speed in healthy older women. This study assessed whether bench-stepping also improves dynamic balance and movement complexity during stair walking.

**Methods:** Stair walking data were obtained from a previous study involving 45 healthy older women (69y±4) that assessed the effects of a 12-week bench-stepping intervention with non-training controls. Centre-of-mass acceleration was measured during stair ascent and descent. Linear dynamics included time, acceleration magnitude, and harmonic ratios (HR; indicating symmetry). Movement complexity was quantified using nonlinear dynamics including sample entropy (SE), recurrence quantification analysis (RQA), and fractal dimension (FD).

**Results:** For stair ascent, increased speed (p =0.018, R^2^_partial_ =0.093,) was accompanied by proportional increases in acceleration magnitudes (p≤0.039, R^2^_partial_ =0.078-0.101). SE decreased more in the intervention group (p ≤0.012, R^2^_partial_ =0.049-0.101), indicating more predictable dynamics. In contrast, for stair descent, no changes in speed or acceleration magnitudes were observed. However, SE (p =0.001, R^2^_partial_ =0.082) and maximum RQA line length (p= 0.008, R^2^_partial_ =0.057) of vertical acceleration increased significantly compared to controls, indicating lower predictability and more persistent recurring patterns. No significant changes were found for other outcomes. Exploratory factor analysis revealed distinct differences in motor behaviour between stair ascent and descent.

**Conclusion:** Bench-stepping training induced measurable changes in stair walking dynamics. Specifically, sample entropy shows potential as a sensitive marker of altered motor complexity, particularly of vertical accelerations. Interestingly, the direction of changes in unpredictability differed between stair ascent and descent, suggesting different underlying control strategies.

## 1. Introduction

Stair walking is a challenging and high-risk activity for older people, placing considerable demands on strength, balance, and coordination [1, 2]. Although falls on stairs are less common than overground falls such as slips and trips, they constitute a disproportionate risk for severe injury and death [3, 4]. Among older people, stair-related falls are the leading cause of in-home fall-related injuries requiring emergency care [5]. Therefore, fall prevention strategies should aim to improve the ability to negotiate stairs safely.

A previous study found that bench-stepping training improved lower-limb power and stair-climbing speed in healthy older women [6]. However, postural sway during balance tasks did not change significantly [5]. This may be attributable to a ceiling effect for improvements due to the relatively fit cohort, limiting transfer effects of the bench-stepping training, or to a lack of sensitivity in the assessment methods to subtle training-related improvements. Although the first explanation cannot be ruled out, more advanced analyses may reveal more sensitive measures and detect improvements in stair walking dynamics.

Linear analyses of kinematic time series have previously been used to assess movement patterns and estimate balance control during stair negotiation. These measures include root mean square of jerk, accelerations and angular velocities, and harmonic ratios, which provide insights into movement smoothness and symmetry [7–11]. However, linear analyses rely on the principle of simple stimulus-response relationships and assume that variability of postural control scales proportionately to the intensity of perturbations during gait [12]. As such, linear measures do not account for the complexity of the interplay among physiological systems underlying movement patterns. In contrast, nonlinear analyses allow for assessments of aspects of motor behaviour like movement complexity, regularity, and adaptability. These nonlinear analyses include entropy-based or fractal-based measures, quantification of recurrence plots, and quantification of stability [11, 12]. Although nonlinear measures are becoming more common in gait analysis [12–14], their application in stair walking remains limited. This may be attributable to the challenging nature of stair walking, especially in older and frail populations, which limits the repetitive performance needed to collect sufficient data. Consequently, nonlinear measures that require a relatively large number of data points or gait cycles, such as those characterising local dynamic stability, are not suitable for analyses of stair walking [15]. However, with proper parameterisation, measures such as sample entropy, fractal dimension, and recurrence quantification analyses can quantify motor behaviour in dynamic tasks with a limited number of movement cycles [12].

The aims of this study were 1) to assess if dynamic balance and movement complexity during stair walking change following bench-stepping training in older women, and 2) to explore potential latent constructs characterising motor behaviour during stair ascent and descent using factor analysis. We hypothesised that 1) linear and nonlinear measures would capture training-related changes in dynamic balance and movement complexity, highlighting these as sensitive markers for changes in stair walking dynamics, and that 2) distinct latent factors of motor behaviour would be found for stair ascent and descent.

### 2. Materials and methods

The data analysed in this study were collected as part of a previous randomised controlled trial, investigating the effects of bench-stepping exercise on muscle volume, strength, and functional performance in healthy older women [6]. Below, a brief overview of the study methodology is provided. For full details regarding recruitment procedures, inclusion and exclusion criteria, ethical approval, and training procedures, we refer to the original study by Baggen et al. [6].

### 2.1. Participants

This study included data from 45 healthy community-living older women (69±4 years). All participants were randomly assigned to the experimental STEEP bench-stepping exercise (n = 24) or non-training control group (n = 21).

### 2.2. Intervention and measurements

In brief, the STEEP intervention consisted of 40 minutes of bench stepping exercise following a progression program with incremental step heights and weighted vests, performed 3 times per week for 12 weeks.

Stair negotiation was assessed pre- and post-training using an inertial sensor (Dynaport MoveTest®, McRoberts, The Hague, NL), attached to the lower back (L4/L5 level) with a Velcro strap. Participants walked up and down a 12-step staircase. Participants were instructed to walk as fast as they felt safe to do, without running or holding the handrails. They were instructed to walk in a foot-over-foot pattern, avoiding placing two feet on the same step or skipping steps. Each stair-walking bout started and ended with three seconds of quiet stance for calibration purposes. All participants performed two bouts of stair ascent and descent pre- and post-intervention.

Inertial data consisted of three-dimensional linear accelerations and angular velocities of the approximate centre-of-mass (CoM), sampled at 100Hz. All data were processed using Matlab (Matlab R2024a, MathWorks®, Natick, MA, USA). Corrections for heading and gravity were applied using a discrete Kalman filter. The filter was initialised using acceleration and gyroscope data from the three seconds of quiet stance before each bout of stair walking. Gravity was reconstructed from estimated roll and pitch and removed from the accelerometer signals. The corrected accelerations were then transformed from the sensor frame to a local navigation frame using a ZYX (yaw–pitch–roll) Euler rotation sequence. No other filters were applied during processing, unless otherwise specified in Table 1. The first and last step from each walking bout were removed before the analyses to avoid distorting effects of start- and end-phase accelerations [8]. To achieve this, heel strikes of the leading foot were approximated from accelerations of the pelvis [16]. Subsequently, each time series was truncated from the first heel strike following gait initiation to the heel strike of the penultimate step. An overview of the measures calculated to characterise stair walking dynamics from the inertial data can be found in Table 1. These variables were included in the analyses as they are robust for use with relatively short time series [12]. Additionally, these measures do not depend on exact step detection, which eliminates a potential source of error [11].

**Table 1.** Overview of gait variables and descriptions.

| Measure | Abbreviation | Description |
| --- | --- | --- |
| Root mean squared acceleration | RMS | Magnitude of Centre-of-mass acceleration in AP, ML, and VT directions separately. Data were filtered using a zero-lag 4<sup>th</sup> order low-pass Butterworth filter at 20Hz. |
| Harmonic ratio | HR | Smoothness and step-to-step symmetry of acceleration in AP, ML, and VT directions. Data were filtered using a zero-lag 4<sup>th</sup> order low-pass Butterworth filter at 20Hz. Acceleration data were segmented into six strides and transformed into the frequency domain using Fourier analysis. Calculated as the ratio of the sum of the amplitudes of even harmonics to odd harmonics. |
| Mean pelvis angular velocity | mPAV | Rotational movement of the pelvis, reflecting postural balance. Calculated using the mean Euclidean magnitude of angular velocity. Results were normalised to walking speed. Data were filtered using a zero-lag 4<sup>th</sup> order low-pass Butterworth filter at 20Hz. |
| Dimensionless jerk | DJ | Smoothness of overall movement, reflecting changes in acceleration over time. The Euclidean magnitude of 3D acceleration was calculated. Instantaneous jerk was obtained by differentiating 3D acceleration with respect to time. The squared jerk was then integrated and normalised to movement duration and acceleration magnitude. |
| Sample entropy | SE | Unpredictability, calculated using acceleration time series in AP, ML and VT directions separately. Data were time-normalised to the average number of data points across all bouts of stair ascent and descent respectively. We used an embedding dimension of 2 and tolerance of 0.2 times the standard deviation and a time delay (Tau) of 1. |
| Recurrence quantification analysis | RQA | Recurrent structure, regularity, and determinism of the time series, performed using acceleration data in AP, ML and VT directions separately. All data were first Z-scored to improve comparability between subjects and conditions, and time-normalised (see Sample Entropy). The most common embedding dimension was determined across trials using the False Nearest Neighbour method (Kennel approach), resulting in $m = 5$. Time delay (Tau) was determined using the minimal mutual information criterion and fixed at 6. In accordance with previous studies, we chose to fix the recurrence rate at 0.05 by using a variable radius. The Theiler window was set at 1, and the minimum diagonal line length at 2. Extracted features were determinism (DET) representing regularity, laminarity (LAM) representing intermittence, mean and maximal line length (Lmean and Lmax) representing persistent recurring patterns. |
| Higuchi fractal dimensions | HFD | Movement complexity, calculated using acceleration data in AP, ML and VT directions separately. The maximum interval parameter (kmax) was set to 8. |

AP = anteroposterior, ML = mediolateral, VT = vertical

### 2.3. Statistical analysis

Statistical analyses of main effects were performed using R (v 4.5.2; R Foundation for Statistical Computing, Vienna, Austria) in RStudio (v2025.09.2; Posit Software, PBC, Boston, MA, USA). To investigate training-related changes, differences in outcomes between groups over time were assessed for each variable using linear mixed-effects models (LMMs). Group (STEEP vs. control), time (pre- and post-intervention), and their interaction (group × time) were included as fixed effects. Subject was included as a random effect to account for between-subject variability and within-subject correlation across timepoints (2 measurements for both assessment times). Models were fitted using restricted maximum likelihood.

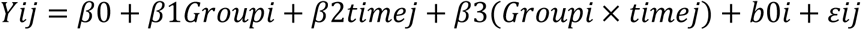

where *b*_0*i*_ is a subject-specific random intercept and *ε_ij_* is the residual error..

Estimated marginal means and corresponding 95% confidence intervals were computed to characterise group differences at each time point and to examine within-group changes over time. Model assumptions were evaluated using simulated residual diagnostics including QQ plots. If assumptions were violated, bootstrap confidence intervals were used to obtain robust estimates of fixed effects. Generalised effect sizes were obtained for interpretation, quantifying the magnitude of fixed effects in terms of mean differences (Cohen’s d) and variance explained (R^2^_partial_) for fixed effects. Significance was set at α = 0.05.

Principal component analysis (PCA) and exploratory factor analysis (EFA) were performed to explore potential latent constructs of stair walking dynamics. Analyses were performed separately for stair ascent and descent because of clear differences in the number of factors extracted. Data were first normalised by Z-scoring and correlations between variables were inspected for redundancy. Subsequently, PCA was used to determine the number of statistically meaningful principal components using the broken-stick method, which informed the number of factors extracted in the subsequent EFA. For EFA, data were first checked for redundancy and congruence of factors. Strong correlations (>0.95) were found between RQA-LAM and RQA-DET, indicating redundancy. However, both were retained as removal significantly lowered congruence (<0.85). Inclusion of Lmax data resulted in significantly lower congruence of factors between pre-and post-measurements. Because Lmax data showed substantial outliers, these data were excluded before pooling. Factor similarity between pre- and post-training loading patterns was assessed using Tucker’s coefficient of congruence. Because congruence exceeded 0.85 for all factors, pre- and post-training data were pooled, and a single model was fitted. Factors were rotated using orthogonal varimax rotation to facilitate interpretability. For each factor, Kaiser’s criterion (eigenvalue > 1) was met, and variables with absolute loading magnitude > 0.5 were considered to meaningfully contribute to that factor [17]. Training-related effects on factor scores were evaluated with a linear mixed-effects model using factor scores, with group, time, and their group × time interaction effects, and a random intercept for participant.

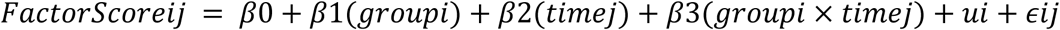

where *u_i_* is the subject-specific random intercept and *ε_ij_* is the residual error.

## 3. Results

Group descriptives can be found in Supplementary Table 1. For detailed data on training effects on muscle volume, strength, power, and functional performance, see Baggen et al. [6]. In brief, bench-stepping exercise significantly increased thigh muscle volume, knee extensor strength and power, and improved functional performance (reduced times for the 5x Sit-to-Stand test, 10m walk test, and stair climbing speed).

### 3.1. Linear measures

Following bench-stepping training, stair ascent time decreased significantly in the STEEP group relative to controls (-0.287 ± 0.062 seconds, β = 0.232, standard error = 0.094, p = 0.018, and R^2^_partial_ = 0.093), indicating faster stair negotiation. This was accompanied by significant increases in RMS of acceleration across all three directions (p ≤ 0.039, R^2^_partial_ = 0.078-0.101), reflecting higher movement amplitudes. No significant changes were observed for harmonic ratios, mean pelvis angular velocity, or dimensionless jerk during ascent (p > 0.05). For stair descent, no significant changes were found for any linear measure (p > 0.05).

### 3.2. Nonlinear measures

SE decreased significantly in the STEEP group for ML and VT directions during stair ascent directions (p = 0.012 and < 0.001, R^2^_partial_ = 0.049 and 0.101 respectively), indicating lower movement unpredictability following training. In contrast, SE increased significantly for the VT direction during stair descent (p = 0.001, R^2^_partial_ = 0.082, reflecting more unpredictable vertical movement. RQA-Lmax increased significantly for the VT direction during descent (p = 0.008, R^2^_partial_ = 0.057), indicating longer maximum duration of recurring vertical movement patterns. No significant changes were found for RQA determinism, laminarity, mean line length, or HFD in either condition (p >0.05). Results are presented in Figure 1 and Supplementary Table 2.

**Figure 1:**
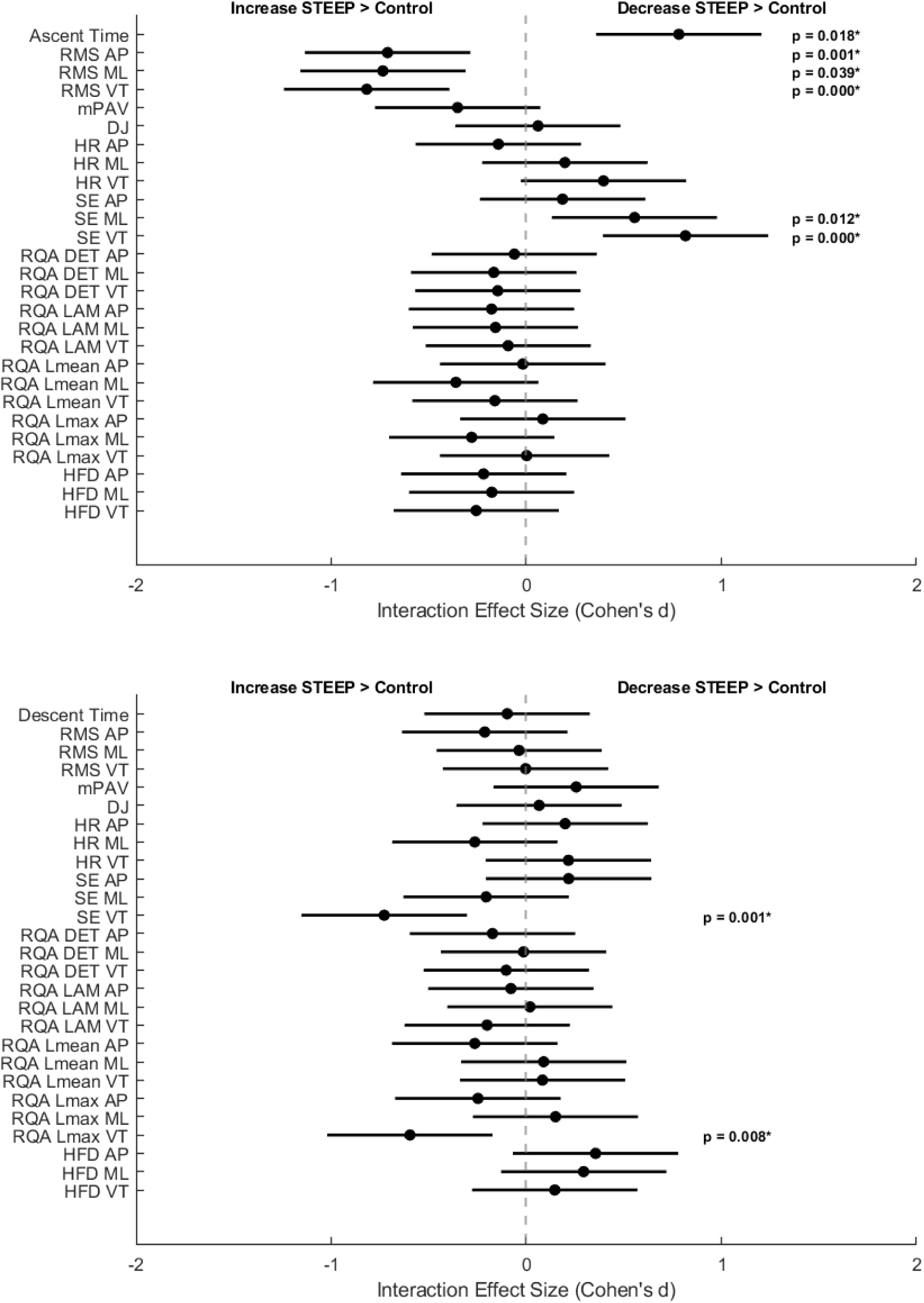
Forest plot group x time interaction effects for stair ascent (Top) and descent (Bottom)

### 3.3. Factor analysis

Following PCA, three components were retained for stair ascent, accounting for 63.9% of the variance, and five components for descent, accounting for 74.3% of the variance (all eigenvalues > 1).

EFA of the three factors for stair ascent showed the strongest loadings in Factor 1 were for ascent time, RMS, SE (ML and VT), RQA-Lmean (ML), and HFD (AP and VT), reflecting a general dimension of movement quality encompassing acceleration magnitude, unpredictability, recurrence patterns, and complexity. Factors 2 and 3 primarily represented movement regularity and complexity in ML and AP directions respectively. Cross-loading occurred for RQA-Lmean ML and HFD AP. Cross-loading was also evident for other factors, but with a clear weight difference between factors. Results are presented in Figure 2 and Supplementary Table 3.

**Figure 2:**
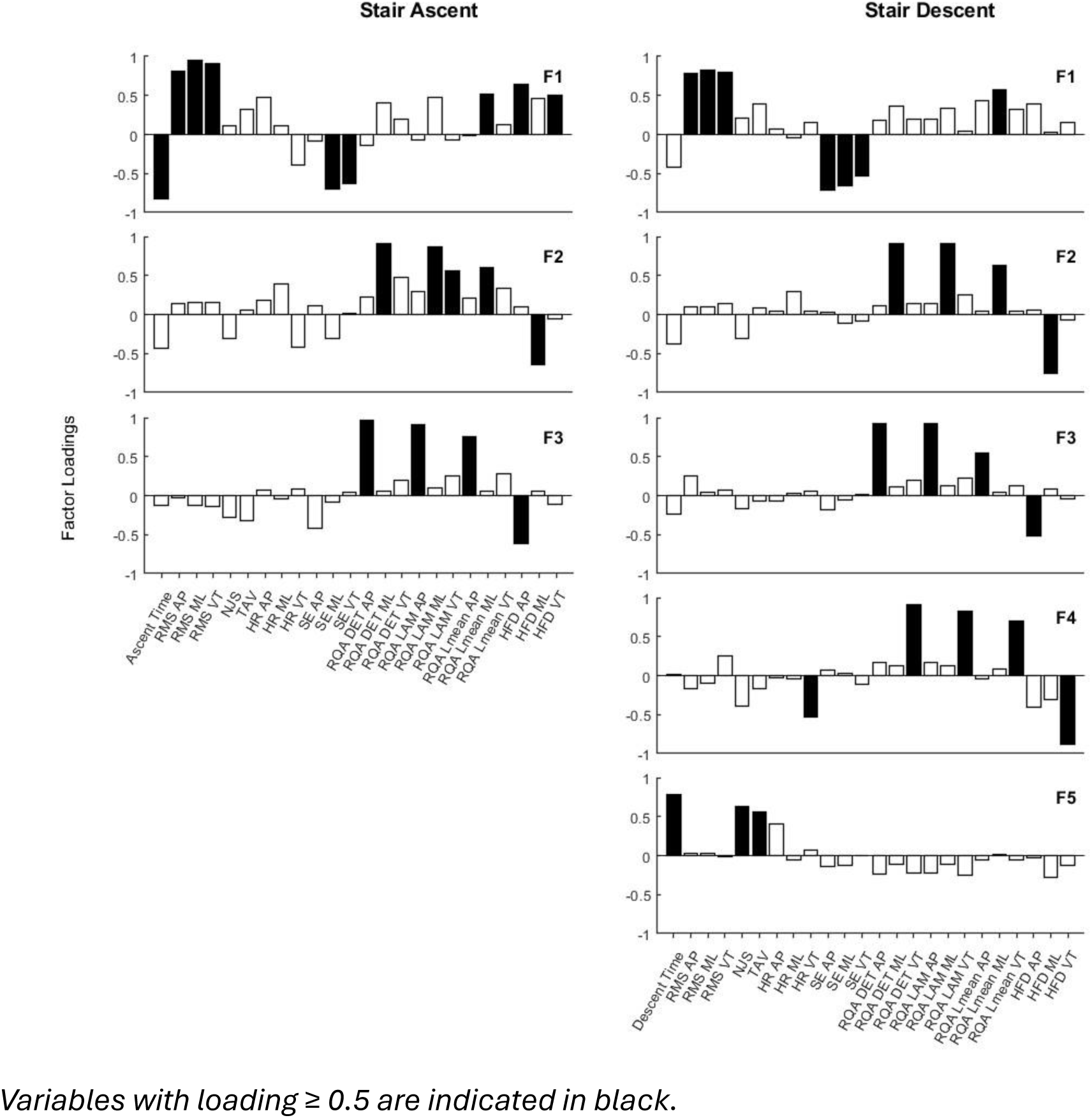
Factors and loadings for stair ascent (Left) and descent (Right).

EFA of the five factors for stair descent showed the strongest loadings in factor 1 in a similar pattern to ascent, except for Time and HFD (AP and VT), which showed higher loadings in different factors. Factors 2 and 3 also represented movement regularity and complexity in ML and AP directions respectively, with an additional factor 4 representing movement regularity and complexity in the VT direction. Factor 5 represented descent time, jerk, and pelvis angular velocity.

Factor 1, extracted from stair ascent, was the only factor to show a significant training-related increase in the factor score (+0.487) compared to the control group (β = -0.54 SE = 0.14, p < 0.001).

**Supplementary Table 1:** Baseline descriptives of the STEEP intervention and control group.

|  | STEEP | CONTROL | mean difference<br>(95% CI) | *p* | Effect<br>size<br>(Cohen<br>'s *d*) |
| --- | --- | --- | --- | --- | --- |
| Age (y) | 69 ± 4 | 69 ± 4 | 0.03 (-2.33 to 2.39) | 0.98 | 0.02 |
| Body mass (kg) | 72 ± 14 | 66 ± 11 | 6.67 (-0.34 to 13.68) | 0.06 | 0.51 |
| Height (cm) | 164 ± 6 | 162 ± 5 | 2.52 (-0.88 to 5.91) | 0.14 | 0.36 |
| BMI (kg/m<sup>2</sup>) | 26.77 ± 4.70 | 25.09 ± 3.36 | 1.79 (-0.67 to 4.25) | 0.15 | 0.41 |
| Leisure time PA-score | 24.52 ± 24.81 | 22.80 ± 14.53 | 1.26 (-11.00 to 13.51) | 0.84 | 0.08 |
| Handgrip<br>strength (kg) | 28.22 ± 4.77 | 29.00 ± 5.71 | -0.53 (-3.76 to 2.69) | 0.74 | 0.15 |

**Supplementary Table 2:** Group * Time interaction effects table.

| Outcomes | effect estimate<br>control - STEEP | R<sup>2</sup><sub>partial</sub> | df | 95% CI | t | p |
| --- | --- | --- | --- | --- | --- | --- |
| *Time* |  |  |  |  |  |  |
| Ascent | 0.232 | 0.093 | 127.00 | 0.11 to 0.36 | 3.615 | **0.018**†* |
| Descent | -0.046 | 0.002 | 127.00 | -0.24 to 0.15 | -0.453 | 0.651 |
| *Root Mean Square* |  |  |  |  |  |  |
| AP Ascent | -0.033 | 0.078 | 127.00 | -0.05 to -0.01 | -3.29 | **0.001** ** |
| ML Ascent | -0.064 | 0.083 | 127.00 | -0.10 to -0.03 | -3.40 | **0.039**†* |
| VT Ascent | -0.076 | 0.101 | 127.00 | -0.12 to -0.04 | -3.78 | **<0.001** *** |
| AP Descent | -0.005 | 0.008 | 127.00 | -0.01 to 0.00 | -0.99 | 0.524† |
| ML Descent | -0.001 | 0.000 | 127.00 | -0.01 to 0.01 | -0.17 | 0.910† |
| VT Descent | 0.000 | 0.000 | 127.00 | -0.02 to 0.02 | -0.02 | 0.988 |
| *Harmonic ratio* |  |  |  |  |  |  |
| AP Ascent | -0.034 | 0.003 | 127.00 | -0.13 to 0.07 | -0.66 | 0.523† |
| ML Ascent | 0.059 | 0.007 | 127.00 | -0.07 to 0.19 | 0.92 | 0.362 |
| VT Ascent | 0.066 | 0.026 | 127.00 | 0.00 to 0.14 | 1.83 | 0.069 |
| AP Descent | 0.035 | 0.007 | 127.00 | -0.04 to 0.11 | 0.92 | 0.358 |
| ML Descent | -0.057 | 0.012 | 127.00 | -0.15 to 0.03 | -1.22 | 0.225 |
| VT Descent | 0.027 | 0.008 | 127.00 | -0.03 to 0.08 | 1.00 | 0.321 |
| *Mean Pelvis Angular Velocity* |  |  |  |  |  |  |
| Ascent | -0.040 | 0.020 | 127.00 | -0.09 to 0.01 | -1.63 | 0.106 |
| Descent | 0.020 | 0.011 | 127.00 | -0.01 to 0.05 | 1.18 | 0.239 |
| *Dimensionless Jerk* |  |  |  |  |  |  |
| Ascent | 15.980 | 0.001 | 127.00 | -97.55 to 129.51 | 0.28 | 0.783 |
| Descent | 75.650 | 0.001 | 127.00 | -413.40 to 564.70 | 0.30 | 0.839† |
| *Sample Entropy* |  |  |  |  |  |  |
| AP Ascent | 0.014 | 0.006 | 127.00 | -0.02 to 0.05 | 0.86 | 0.391 |
| ML Ascent | 0.046 | 0.049 | 127.00 | 0.01 to 0.08 | 2.57 | **0.012\*** |
| VT Ascent | 0.047 | 0.101 | 127.00 | 0.02 to 0.07 | 3.78 | **<0.001\*\*\*** |
| AP Descent | 0.017 | 0.008 | 127.00 | -0.02 to 0.05 | 1.00 | 0.318 |
| ML Descent | -0.011 | 0.007 | 127.00 | -0.03 to 0.01 | -0.95 | 0.342 |
| VT Descent | -0.031 | 0.082 | 127.00 | -0.05 to -0.01 | -3.37 | **0.001\*\*** |
| *Recurrence Quantification Analysis (Determinism)* |  |  |  |  |  |  |
| AP Ascent | -0.004 | 0.001 | 127.00 | -0.04 to 0.03 | -0.28 | 0.770 |
| ML Ascent | -0.010 | 0.005 | 127.00 | -0.04 to 0.02 | -0.77 | 0.579† |
| VT Ascent | -0.007 | 0.004 | 127.00 | -0.03 to 0.01 | -0.67 | 0.501 |
| AP Descent | -0.011 | 0.005 | 127.00 | -0.04 to 0.02 | -0.80 | 0.425 |
| ML Descent | -0.001 | 0.000 | 127.00 | -0.03 to 0.02 | -0.07 | 0.948 |
| VT Descent | -0.002 | 0.002 | 127.00 | -0.01 to 0.01 | -0.48 | 0.636 |
| *Recurrence Quantification Analysis (Laminarity)* |  |  |  |  |  |  |
| AP Ascent | -0.009 | 0.005 | 127.00 | -0.03 to 0.01 | -0.82 | 0.412 |
| ML Ascent | -0.006 | 0.004 | 127.00 | -0.02 to 0.01 | -0.73 | 0.600† |
| VT Ascent | -0.003 | 0.001 | 127.00 | -0.02 to 0.01 | -0.43 | 0.669 |
| AP Descent | -0.003 | 0.001 | 127.00 | -0.02 to 0.01 | -0.37 | 0.715 |
| ML Descent | 0.001 | 0.000 | 127.00 | -0.01 to 0.01 | 0.09 | 0.931 |
| VT Descent | -0.003 | 0.007 | 127.00 | -0.01 to 0.00 | -0.93 | 0.355 |
| *Recurrence Quantification Analysis (Mean line length)* |  |  |  |  |  |  |
| AP Ascent | -0.006 | 0.000 | 127.00 | -0.13 to 0.12 | -0.08 | 0.933 |
| ML Ascent | -0.253 | 0.022 | 127.00 | -0.55 to 0.04 | -1.67 | 0.097 |
| VT Ascent | -0.078 | 0.004 | 127.00 | -0.28 to 0.13 | -0.74 | 0.458 |
| AP Descent | -0.080 | 0.012 | 127.00 | -0.21 to 0.05 | -1.22 | 0.224 |
| ML Descent | 0.041 | 0.001 | 127.00 | -0.15 to 0.23 | 0.41 | 0.680 |
| VT Descent | 0.044 | 0.001 | 127.00 | -0.18 to 0.27 | 0.39 | 0.700 |
| *Recurrence Quantification Analysis (Max line length)* |  |  |  |  |  |  |
| AP Ascent | 1.882 | 0.001 | 127.00 | -7.50 to 11.26 | 0.39 | 0.773† |
| ML Ascent | -14.797 | 0.013 | 127.00 | -37.23 to 7.64 | -1.29 | 0.318† |
| VT Ascent | 0.115 | 0.000 | 127.00 | -25.31 to 25.54 | 0.01 | 0.993† |
| AP Descent | -5.596 | 0.010 | 127.00 | -15.15 to 3.96 | -1.15 | 0.348† |
| ML Descent | 11.080 | 0.004 | 127.00 | -20.29 to 42.46 | 0.69 | 0.558† |
| VT Descent | -42.960 | 0.057 | 127.00 | -73.49 to -12.43 | -2.76 | **0.008†\*\*** |
| *Higuchi Fractal Dimensions* |  |  |  |  |  |  |
| AP Ascent | -0.012 | 0.008 | 127.00 | -0.04 to 0.01 | -1.01 | 0.314 |
| ML Ascent | -0.007 | 0.005 | 127.00 | -0.02 to 0.01 | -0.82 | 0.414 |
| VT Ascent | -0.013 | 0.011 | 127.00 | -0.03 to 0.01 | -1.19 | 0.238 |
| AP Descent | 0.024 | 0.021 | 127.00 | 0.00 to 0.05 | 1.64 | 0.103 |
| ML Descent | 0.013 | 0.014 | 127.00 | -0.01 to 0.03 | 1.36 | 0.176 |
| VT Descent | 0.007 | 0.004 | 127.00 | -0.01 to 0.03 | 0.68 | 0.500 |

Positive effect estimates indicate a greater decrease in STEEP vs Control, negative effect estimates indicate a greater increase in STEEP vs Control. \*, \*\*, \*\*\* = significant group\*time interaction effect at $\alpha = 0.05, 0.01$ , and $0.001$ respectively, † = reported p from fixed-effect coefficients.

**Supplementary Table 3:** Results of exploratory factor analysis.

| Variable | Ascent |  |  | Descent |  |  |  |  |
| --- | --- | --- | --- | --- | --- | --- | --- | --- |
|  | Factor 1 | Factor 2 | Factor 3 | Factor 1 | Factor 2 | Factor 3 | Factor 4 | Factor 5 |
| Time | **-0.831** | -0.428 |  | -0.417 |  |  |  | **0.789** |
| RMS AP | **0.809** |  |  | **0.781** |  |  |  |  |
| RMS ML | **0.944** |  |  | **0.822** |  |  |  |  |
| RMS VT | **0.916** |  |  | **0.803** |  |  |  |  |
| DJ |  |  |  |  |  |  |  | **0.625** |
| mPAV |  |  |  |  |  |  |  | **0.565** |
| HR AP | 0.479 |  |  |  |  |  |  | 0.406 |
| HR ML |  |  |  |  |  |  |  |  |
| HR VT |  | -0.421 |  |  |  |  | **-0.530** |  |
| SE AP |  |  | -0.426 | **-0.716** |  |  |  |  |
| SE ML | **-0.693** |  |  | **-0.661** |  |  |  |  |
| SE VT | **-0.628** |  |  | **-0.532** |  |  |  |  |
| RQA DET AP |  |  | **0.961** |  |  | **0.930** |  |  |
| RQA DET ML | 0.406 | **0.909** |  |  | **0.910** |  |  |  |
| RQA DET VT |  | 0.482 |  |  |  |  | **0.909** |  |
| RQA LAM AP |  |  | **0.915** |  |  | **0.918** |  |  |
| RQA LAM ML | 0.477 | **0.862** |  |  | **0.912** |  |  |  |
| RQA LAM VT |  | **0.562** |  |  |  |  | **0.824** |  |
| RQA Lmean AP |  |  | **0.752** | 0.441 |  | **0.540** |  |  |
| RQA Lmean ML | **0.516** | **0.597** |  | **0.571** | **0.626** |  |  |  |
| RQA Lmean VT |  |  |  |  |  |  | **0.699** |  |
| HFD AP | **0.647** |  | **-0.619** |  |  | **-0.517** | -0.403 |  |
| HFD ML | 0.464 | **-0.644** |  |  | **-0.752** |  |  |  |
| HFD VT | **0.501** |  |  |  |  |  | **-0.888** |  |
| p interaction | <0.001* | 0.436 | 0.386 | 0.628 | 0.945 | 0.395 | 0.581 | 0.444 |

Loading of Z-scored variables per factor extracted separately for stair ascent and descent. Absolute loadings $\geq 0.4$ are shown. Loadings $\geq 0.5$ in bold. p-values of LMM interaction effects are displayed below each factor. \* = significant interaction effect.

## 4. Discussion

This analysis suggest that a 12-week bench-stepping intervention in older women induced measurable, task-specific changes in stair walking dynamics. In addition to previously reported changes in ascent speed [6], linear measures showed increased RMS during stair ascent, while non-linear measures revealed changes in sample entropy during both ascent and descent, and in RQA Lmax during stair descent. Sample entropy was the most sensitive metric, identifying training-related alterations in stair-walking dynamics that were not captured by most conventional or other nonlinear measures. The direction of these changes differed between stair ascent and descent, suggesting that training influenced motor behaviour differently across the two tasks. Exploratory factor analysis also indicated that stair ascent and descent are characterised by partly distinct underlying dimensions of movement control.

### 4.1. Linear outcomes

Bench-stepping training improved stair climbing performance relative to controls. The decrease in stair ascent time indicates improved functional performance, with corresponding increases in magnitude of acceleration representing the greater movement impulses associated with faster stair ascent. Previous research in people with multiple sclerosis has linked increases in RMS to decreased stability [18], suggesting a gain of stair ascent speed at the cost of dynamic stability in the intervention group. However, adding walking time as a factor post-hoc resulted in no additional interaction effects on RMS, indicating that the magnitude of CoM accelerations changed proportionally to stair ascent speed. Therefore, the increased RMS accelerations during stair ascent in this study likely did reflect an increase in speed and not a change in dynamic stability. Training effects on stair descent appeared to be limited, with no significant changes in any linear outcomes.

### 4.2. Nonlinear outcomes

For stair ascent, SE of ML and VT accelerations was reduced following bench-stepping training, indicating more predictable acceleration patterns following bench-stepping training, despite increases in stair climbing speed and magnitude. In the ML direction, lower sample entropy suggested fewer corrections in lateral movement. ML control is a challenging aspect of stair walking due to inherent gait instability and high sagittal axis angular momentum in this plane during stair ascent [19–21]. Lower sample entropy in the VT direction indicates more regular stepping dynamics, which suggests better control of the loading and unloading phases, more controlled transfer of body mass between steps, and enhanced intersegmental coordination of the lower limbs and trunk. Sample entropy is known to be sensitive to gait speed, specifically due to differences in vector lengths [22]. Therefore, all acceleration data were time-normalised before calculating time-sensitive metrics such as SE and RQA. However, this does not fully negate potential confounding effects of frequency-related changes in rhythmicity. Nevertheless, if changes in rhythmicity were the predominant cause for lowered entropy, this would be reflected in concurrent increases in harmonic ratios. Although there is some evidence of training-related increases in HR during stair ascent, these changes were not significant. Therefore, we conclude that the more predictable movement patterns indicate training-related improvements during stair ascent.

For stair descent, despite the absence of training-related changes in descent time or movement amplitude, subtle changes were observed in vertical movement control and temporal organisation of gait dynamics. In contrast with stair ascent, SE-VT during stair descent increased significantly. These results may appear contradictory. However, entropy analyses (like most nonlinear metrics of movement complexity) operate with the assumption of an optimal window of movement variability, rather than depending on normative values [23]. Additional comparisons of baseline SE-VT in the intervention group revealed that the SE for ascent (0.64) was around the average among all SE scores obtained from both ascent and descent in this study (0.65), whereas the SE for descent was lower (0.48). Therefore, it is reasonable to assume that the higher entropy values, in the absence of significant changes in stair descent time or movement magnitude, may suggest greater adaptability, with increased Lmax indicating improved stability.

The limited changes in motor behaviour during stair descent compared to ascent may be attributable to a lower task-specificity of the bench-stepping exercises, as participants only performed backward and sideways step-downs during training, which may have provided limited transfer to forward stair descent. This is particularly relevant given the importance of task specificity in ageing populations [24, 25]. The primary purpose of the bench-stepping intervention was to improve concentric lower limb muscle strength and power, which are limiting factors in stair ascent [6, 26]. In contrast, during stair descent, eccentric strength and balance control are more important [2, 27]. Although eccentric muscle strength could have improved from backwards step-downs, it was not tested. Nevertheless, the lack of balance improvements reported previously [6] indicates that potential transfer effects and concurrent changes in motor patterns were likely small. These findings suggest that rehabilitation programs targeting stair descent safety should incorporate descent-specific training.

### 4.3. Factor composition

In line with the differences in effects of bench-stepping exercise on movement unpredictability during stair ascent and descent, exploratory factor analysis revealed differences between the two tasks. Although stair ascent and descent did show some similarities in factor composition, the greater number of extracted factors for descent suggests that additional motor control processes are employed during stair descent. The most prominent differences were the presence of two additional factors (i.e. Factor 4 and 5) for stair descent, one with predominant loading by VT parameters, which may represent control of CoM deceleration, and another representing speed. During stair ascent, speed is primarily determined by the ability of the lower-extremity muscles to generate positive mechanical work to raise the CoM against gravity. In contrast, stair descent is less constrained and involves dissipation and control of gravitational acceleration, placing greater demands on balance control. Additionally, a trip during stair descent is more likely to result in an injurious fall [28], increasing concerns about falling. These factors may elicit tighter and potentially more cognitive control over movement [29]. Overall, the greater number of factors and occurrence of distinct direction-based factors confirms our second hypothesis. Interestingly, the dominant source of shared variance for some factors was the direction of acceleration, rather than the applied metric. This indicates that selection of the most relevant direction of acceleration may be more important than the nonlinear metric used to quantify movement complexity. Finally, only one factor for stair ascent revealed significant training-related effects. This factor represented a general dimension of movement quality, containing particularly high loadings for ascent time, acceleration magnitude, unpredictability, as well as complexity. The increased factor score for the intervention group suggests that participants adopted a faster, more vigorous movement strategy accompanied by altered complexity characteristics of CoM accelerations. As such, the measures represented by this factor should be prioritised when quantifying stair walking dynamics in future studies.

### 4.4. Methodological considerations and limitations

Although significant training-related differences were found in the present study, the inclusion of only healthy post-menopausal women has to be taken into account when interpreting these results. Furthermore, participants generally had a high level of fitness, indicated by high scores on the short physical performance battery and Godin leisure-time exercise questionnaire [6]. This may have introduced a ceiling effect in physical performance, consequently limiting the potential to induce changes in movement complexity. Furthermore, due to the exploratory nature of this study, analyses were aimed at identifying sensitivity and potentially relevant associations across individual linear and nonlinear variables. Therefore, unadjusted p-values were reported to maximise sensitivity to potentially meaningful interaction effects. Results from this study are intended as hypothesis-generating, rather than confirmatory. In addition, although factor analysis provided insight into latent structure, these constructs are statistical representations and do not directly reflect underlying physiological mechanisms. Further work combining neuromuscular and neurophysiological measures is needed to better interpret these control processes. Finally, although the effects of ageing on nonlinear parameters of normal gait have been reported before [14], no evidence is available regarding age-related changes in stair negotiation dynamics. As such, future studies should aim to include more physically frail and sedentary populations, include comparisons between age groups, and focus on specific nonlinear variables and movement directions that have shown potential to detect subtle changes, such as sample entropy and ML accelerations.

### 4.5. Research and clinical implications

Stair walking is among the most physically demanding and fall-prone activities in daily life for older adults, yet rehabilitation programs rarely distinguish between ascent and descent as separate targets. This study highlights that stair ascent and descent impose distinct control demands, and should not be treated as interchangeable phases of the same task. The primary task during ascent is to generate positive work to elevate the CoM against gravity, whereas descent involves controlling the CoM’s gravitational acceleration through eccentric muscle action and negative work [30]. This is reflected by differences in joint kinematics and kinetics, with greater hip and knee angles and moments during ascent, and greater ankle dorsi- and plantarflexion during descent [31]. Treating stair negotiation as a single outcome may therefore obscure meaningful task-specific changes in motor control and lead to exercise programs that incompletely address the demands of both directions.

Nonlinear measures, particularly sample entropy, provided complementary sensitivity to these task-specific adaptations, capturing changes in movement organisation that were not consistently detected by linear measures. Clinically, these findings suggest that improvements in stair performance should not be interpreted as uniform changes in locomotor control. Individuals may exhibit similar gains in overall stair ability while adapting differently in ascent-versus descent-specific control processes. This is particularly relevant for fall prevention, as descent places higher demands on eccentric control and balance regulation and is more strongly associated with injurious falls [28]. Rehabilitation programs targeting stair safety should therefore include descent-specific components, such as forward step-down practice and eccentric loading progressions, rather than assuming that ascent-focused training will generalise. The clinical utility of nonlinear kinematic measures such as sample entropy will ultimately depend on establishing their minimal detectable change and linking them to patient-relevant outcomes including falls and stair-related balance confidence. As wearable inertial sensors become more accessible in clinical settings, these measures hold promise as sensitive tools for monitoring motor control quality during rehabilitation, complementing conventional assessments of speed and step performance.

### 4.6. Conclusion

This study provides evidence that nonlinear measures can be used as markers for training-related changes in movement complexity. Among the nonlinear measures examined, sample entropy has potential as a sensitive marker for training-related changes, particularly for vertical accelerations. However, the direction of changes in unpredictability differed between stair ascent and descent, which represent distinct locomotor constructs, characterised by different patterns of control and task-specific adaptations following training. These findings highlight that movement complexity should be interpreted within the context of task demands. Overall, these findings support the use of nonlinear measures as complementary tools for characterising motor behaviour, but emphasise the importance of distinguishing between stair ascent and descent as separate constructs in both research and clinical assessment.

## Data Availability

All data produced in the present study are available upon reasonable request to the authors

